# Performance metrics for models designed to predict treatment effect

**DOI:** 10.1101/2022.06.14.22276387

**Authors:** C.C.H.M. Maas, D.M. Kent, M.C. Hughes, R. Dekker, H.F. Lingsma, D. van Klaveren

**Author notes:** **Materials & Correspondence** C.C.H.M. Maas, Erasmus University Medical Center, Doctor Molewaterplein 40, 3015 GD Rotterdam, Netherlands.

## Abstract

**Background:** Measuring the performance of models that predict individualized treatment effect is challenging because the outcomes of two alternative treatments are inherently unobservable in one patient. The C-for-benefit was proposed to measure discriminative ability. However, measures of calibration and overall performance are still lacking. We aimed to propose metrics of calibration and overall performance for models predicting treatment effect.

**Methods:** Similar to the previously proposed C-for-benefit, we defined observed pairwise treatment effect as the difference between outcomes in pairs of matched patients with different treatment assignment. We redefined the E-statistics, the cross-entropy, and the Brier score into metrics for measuring a model’s ability to predict treatment effect. In a simulation study, the metric values of deliberately “perturbed models” were compared to those of the data-generating model, i.e., “optimal model”. To illustrate these performance metrics, different modeling approaches for predicting treatment effect are applied to the data of the Diabetes Prevention Program: 1) a risk modelling approach with restricted cubic splines; 2) an effect modelling approach including penalized treatment interactions; and 3) the causal forest.

**Results:** As desired, performance metric values of “perturbed models” were consistently worse than those of the “optimal model” (E_avg_-for-benefit≥0.070 versus 0.001, E_90_-for-benefit≥0.115 versus 0.003, cross-entropy-for-benefit≥0.757 versus 0.733, Brier-for-benefit≥0.215 versus 0.212). Calibration, discriminative ability, and overall performance of three different models were similar in the case study. The proposed metrics were implemented in a publicly available R-package “HTEPredictionMetrics”.

**Conclusion:** The proposed metrics are useful to assess the calibration and overall performance of models predicting treatment effect.

## 1. Introduction

Clinicians and patients generally select the treatment that is expected to be beneficial on average for the patient population. However, the average treatment effect (ATE) for a population does not accurately reflect the effect of treatment for each patient individually[1-3]. Various models have been proposed for predicting individualized treatment effects[4]. These models aim to predict the difference between the outcomes of two alternative treatments for each patient.

Usually, only one of the outcomes can be observed for a given patient, the counterfactual outcome remains unobserved. This phenomenon–known as the fundamental problem of causal inference–complicates the assessment of a model’s ability to predict treatment effect. As a result, the performance of models that predict treatment effect cannot be quantified with conventional metrics evaluating risk predictions[5]. To resolve this issue, observed pairwise treatment effect can be defined as the difference between outcomes in pairs of matched patients. Recently, the C-for-benefit has been proposed for quantifying to what extent the models can discriminate between patients who benefit and those who do not[6]. However, measures of calibration–the agreement between predicted and observed treatment effect in *groups* of patients–and measures of overall performance–the discrepancy between predicted and observed treatment effect across *individual* patients–are still lacking.

For models predicting outcome risk and not treatment effect, several metrics are available to assess calibration (i.e., E-statistic), and overall performance (i.e., cross-entropy and Brier score)[7-9]. However, these metrics may poorly reflect a model’s ability to predict treatment effect. For example, in a simulation scenario with a relatively small simulated data sample, the risk predictions of a model with all possible treatment interactions are reasonably well calibrated (Figure 1A), while the corresponding treatment effect predictions are poorly calibrated (Figure 1B)[10]. Apart from such graphical assessment of calibration in groups of patients with similar predicted treatment effects, no metrics are available that quantify the calibration or the overall performance of treatment effect predictions[11].

**Figure 1.**
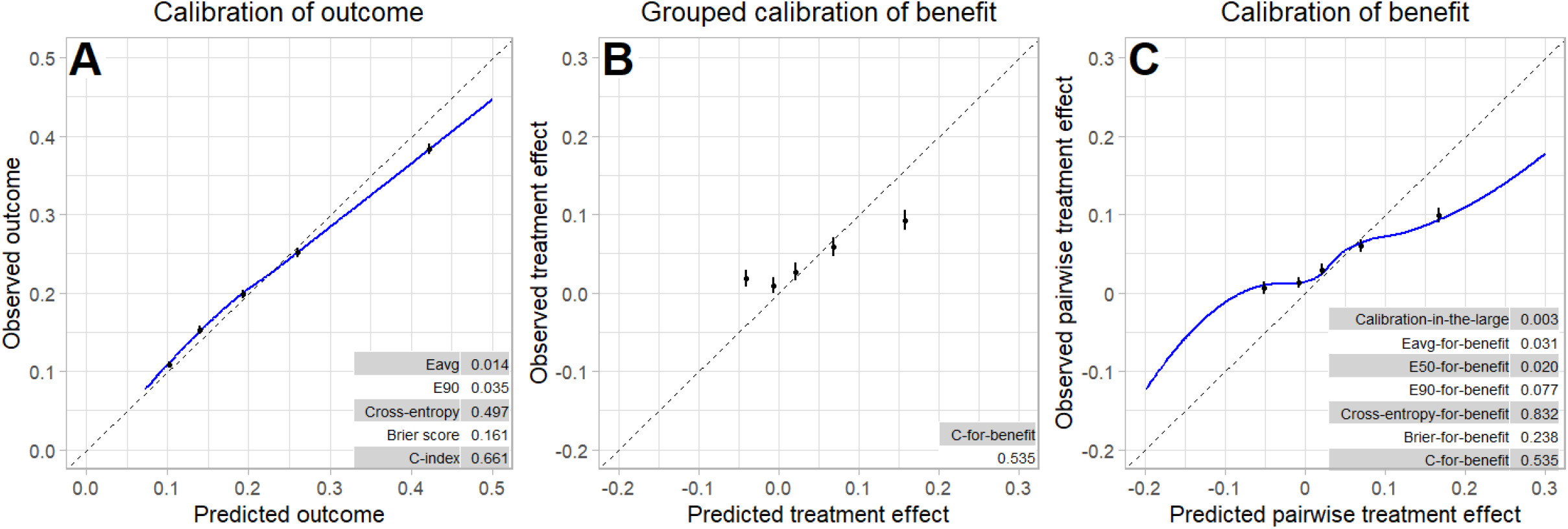
Illustration of risk and benefit calibration figures with performance metrics of simulated data. We sampled (n=3,600) from a simulated trial super population (100,000) with 12 binary risk predictors with 6 true treatment interactions[10]. Panel A depicts observed outcome versus predicted outcome by local regression (blue line, displayed between 0 and 0.5) and quantiles of predicted outcome (black dots), with the E-statistics, cross-entropy, Brier score, and C-index. Panel B depicts the calibration for benefit in groups with confidence intervals, with the C-for-benefit. Panel C depicts observed versus predicted pairwise treatment effect by local regression (blue lane, displayed between -0.2 and 0.3) and quantiles of predicted pairwise treatment effect (black dots), with the newly proposed metrics.

Therefore, we aimed to extend these performance metrics for calibration and overall performance for risk prediction models to models that are designed to predict treatment effect.

## 2. Methods

### 2.1 Definition of treatment effect

With the potential outcomes framework, we can define the (conditional average) treatment effect *τ(x)* for a patient with baseline characteristics *x* as the expected difference between the outcome under control treatment *Y*_*i*_ (0) and the outcome under active treatment *Y*_*i*_(1), conditional on the patient characteristics *x*, i.e.

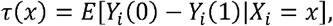

where the potential outcomes *Y*_*i*_ (*W*_*i*_) indicate the outcome *Y*_*i*_ conditional on treatment *W*_*i*_[12]. Here, the event associated with the outcome was assumed to be unfavorable. Thus, treatment benefit, i.e., a positive *τ*(*x*), is expected when the outcome probability under control treatment is higher than the outcome probability under active treatment. Alternatively, two active treatments can be administered instead of control and active treatment.

### 2.2 Metrics based on the matching principle

Using the matching principle, we defined observed pairwise treatment effect as the difference in outcomes between two similar patients with different treatment assignments (Additional file 1)[6]. Similarity was based on baseline patient characteristics to create pairs of similar patients with different treatment assignments. Specifically, we matched each untreated patient with the nearest treated patient based on the Mahalanobis distance between the patient characteristics without replacement[13]. With a binary outcome (say, 0 for alive and 1 for dead), four outcome combinations are possible for a pair of patients. First, treatment benefit was indicated if the treated patient lives and the untreated patient dies. Second, treatment harm was indicated if the treated patient dies and the untreated patient lives. Lastly, no effect of treatment was indicated if both the treated and untreated patients live, or if both die. Thus, the observed pairwise treatment effect takes the values 1 (benefit), 0 (no effect), and -1 (harm). Concurrently, predicted pairwise treatment effect is the difference between the predicted outcome probability of the untreated patient minus the predicted outcome probability of the treated patient (Additional file 1). All of the following metrics use this matching principle and are added to Figure 1C for illustration.

#### 2.2.1 Calibration

Calibration refers to the correspondence between the predicted and observed treatment effects. The calibration-in-the-large or mean calibration was defined as the average observed pairwise treatment effect minus the average predicted pairwise treatment effect[14]. If the algorithm overestimates treatment effect, the average predicted pairwise treatment effect is higher than the observed pairwise treatment effect, resulting in a negative calibration-in-the-large value. Conversely, the calibration-in-the-large will be positive if treatment effect is underestimated.

Calibration can also be assessed by a smoothed calibration curve obtained by a local regression of observed pairwise treatment effect on predicted pairwise treatment effect, with default values for the span and the degree of polynomials (Figure 1C). Similar to the E-statistic and the Integrated Calibration Index (ICI), we propose to measure calibration by the average absolute vertical distance between this smoothed calibration curve and the diagonal line of perfect calibration[7]. This quantity, which we named the E_avg_-for-benefit, can be interpreted as the weighted difference between observed pairwise treatment effect and predicted pairwise treatment effect, with weights determined by the empirical density function of the predicted pairwise treatment effect. Similarly, we defined the E_50_-for-benefit and the E_90_-for-benefit as the median and 90^th^ percentile of the absolute differences between the predicted pairwise treatment effect and the smoothed observed pairwise treatment effect (Additional file 1)[7]. Thus, the E-statistics indicate perfect calibration when zero.

#### 2.2.2 Discrimination

Discrimination refers to a model’s ability to separate patients who benefit from treatment and those who do not. To measure discrimination, we used the previously proposed C-for-benefit, i.e., the probability that from two randomly chosen matched patient pairs with unequal observed pairwise treatment effect, the pair with greater observed pairwise treatment effect also has a larger predicted pairwise treatment effect[6]. The C-for-benefit was calculated by the number of concordant pairs divided by the number of concordant and discordant pairs. Two patient pairs are concordant if the pair with the larger observed pairwise treatment effect also has a larger predicted pairwise treatment effect. Two patient pairs are discordant if the pair with larger observed benefit has a smaller predicted pairwise treatment effect. Two patient pairs are uninformative if the pairs have the same observed pairwise treatment effect. The C-for-benefit is 0.5 if the model cannot distinguish between patients any better than random treatment assignment, and 1 if the model can perfectly distinguish between patients who benefit from treatment and who do not.

#### 2.2.3 Overall performance measures

We propose to measure overall performance using the multi-class versions of the Brier score and cross-entropy because observed pairwise treatment effect can belong to one of three classes (benefit, no effect, harm)[8, 9]. We defined cross-entropy-for-benefit as the logarithmic distance between predicted and observed pairwise treatment effect and Brier-for-benefit as the average squared distance between predicted and observed pairwise treatment effect (Additional file 2). Thus, the overall performance metrics indicate better optimal performance when closer to zero. The cross-entropy-for-benefit and Brier-for-benefit measure overall model performance since these metrics are affected by calibration and discrimination simultaneously. The proposed metrics were implemented in a publicly available R-package “HTEPredictionMetrics”[15].

### 2.3 Data

To illustrate the proposed metrics, we used data from the Diabetes Prevention Program (DPP). The participants of DPP were at risk to develop diabetes, which is defined as a body mass index of 24 or higher and impaired glucose metabolism[16]. The participants were randomized between 1996 and 2001 to receive 1) an intensive program of lifestyle modification lessons, 2) 850 mg of metformin twice a day and standard lifestyle modification, or 3) placebo twice a day and standard lifestyle recommendations. To predict the effect of the intervention on the outcome, i.e., the risk of developing diabetes, we used the patient characteristics sex, age, ethnicity, body mass index, smoking status, fasting blood sugar, triglycerides, hemoglobin, self-reported history of hypertension, family history of diabetes, self-reported history of high blood glucose, and gestational diabetes mellitus (Additional file 3). We imputed missing values of patient characteristics using Multivariate Imputations by Chained Equations[17].

### 2.4 Simulation study

We simulated the outcomes of the DPP using the patient characteristics to study if the proposed performance metrics were better for the model used for outcome generation (“optimal model”) than for deliberately “perturbed models”. The “optimal model” was a logistic regression to model the probability of the outcome (developing diabetes) *p*_*i*_ based on the treatment (e.g., lifestyle intervention) assignment indicator *W*, a centered prognostic index *PI*, and their interaction:

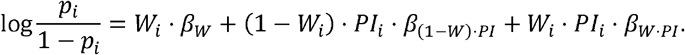

The prognostic index 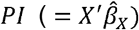 was determined by regressing the outcome variable on the patient characteristics.

Next, we created a super population by duplicating the matched patient pairs 500 times to obtain high precision to ensure that observed differences between metrics are “true” differences. The outcomes of the super population *Y*_*i*_ were simulated from a Bernoulli distribution with the outcome probabilities *p*_*i*_ generated by the “optimal model”.

We then created three deliberate perturbations of the “optimal model”. The first “perturbed model” overestimates ATE by multiplying the coefficient of the treatment assignment indicator (*β*_*W*_) with 2 (Additional file 4; 5). The second “perturbed model” overestimates risk heterogeneity by multiplying the coefficient of the prognostic index for both control treatment(*β*(_*1-W*)·*PI*_) and active treatment (*β*_*W·PI*_) by 2 (Additional file 4; 5). The third “perturbed model” overestimates treatment effect heterogeneity by multiplying the coefficient of the prognostic index of control treatment (*β*_(1*-W*)·*PI*_) by 2 and the coefficient of the prognostic index of active treatment (*β*_*W·PI*_) by 0.5 (Additional file 4; 5). We calculated the root mean squared error (RMSE) to indicate the level of perturbation for each model.

Finally, we computed the performance metrics for the “optimal” and the three “perturbed models” in the super population. We also visualized the performance of each of the four models with treatment effect calibration plots.

### 2.5 Case study

The performance of three different modelling approaches to predict treatment effect for patients at risk of diabetes in the DPP data set was compared using the proposed metrics.

The first approach (“risk model”) uses logistic regression to explain the outcome probability *p*_*i*_ *= P*(*Y*_*i*_ *= 1*|*x*_*i*_ *= x, W*_*i*_ *= w*) based on the treatment indicator *W*, the centered prognostic index *PI* as defined before, and their interaction:

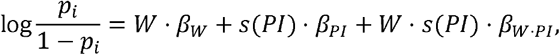

where *s*(·) represents restricted cubic splines with two degrees of freedom.

The second approach (“effect model”) uses a penalized Ridge logistic regression to explain the outcome probability *p*_*i*_ based on the unpenalized treatment indicator *W*, penalized patient characteristics *x*, and their interaction:

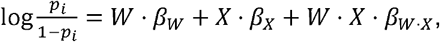

where the level of penalization was determined by the minimum squared error in 5-fold cross-validation[18].

The third approach is a causal forest, which is similar to a random forest but maximizes heterogeneity in treatment effect rather than variation in the outcome[19]. Causal trees were built honestly by partitioning the data into two subsamples. One subsample was used to construct the trees, and another subsample to predict the treatment effect[19]. We used 1000 trees to tune the parameters (e.g., minimal node size, pruning) and 2000 trees to construct the final causal forest.

The models were trained on 70 percent of the patient data. The remaining 30 percent of the patient data, the test set, was used to calculate performance metrics with confidence intervals using 100 bootstrap samples of matched patient pairs. We used the R packages MatchIt for matching patients, mice for single imputation, stats for local regression, rms for restricted cubic splines, glmnet for Ridge penalization, and grf for causal forest (R version 4.1.0)[17, 20-24].

## 3. Results

### 3.1 Patient data

Between 1996 and 2001, the DPP collected data on 3,081 participants of which 1,024 received lifestyle intervention, 1,027 received metformin, and 1,030 received placebo treatment (Additional file 3). The median age of the participants was 52 years (IQR: 42-57 years), 67% of the participants were female, and the median BMI value was 33 (IQR: 29-37). The proportion of patients developing diabetes was 4.8%, 7.0%, and 9.5% among participants receiving lifestyle intervention, metformin, and placebo treatment, respectively (Additional file 3).

### 3.2 Simulation study

As expected, the treatment effect predictions of the “optimal model” were almost perfectly calibrated (calibration-in-the-large=-0.001, E_avg_-for-benefit=0.001, E_50_-for-benefit=0.001, E_90_-for-benefit=0.003, Figure 2A). The “optimal model” was well able to discriminate (C-for-benefit=0.655, Figure 2A) between patients with small treatment harm (ATE=-0.023 in the quantile of patients with smallest predicted pairwise treatment effect) and patients with substantial treatment benefit (ATE=0.385 in the quantile of patients with largest treatment effect).

**Figure 2.**
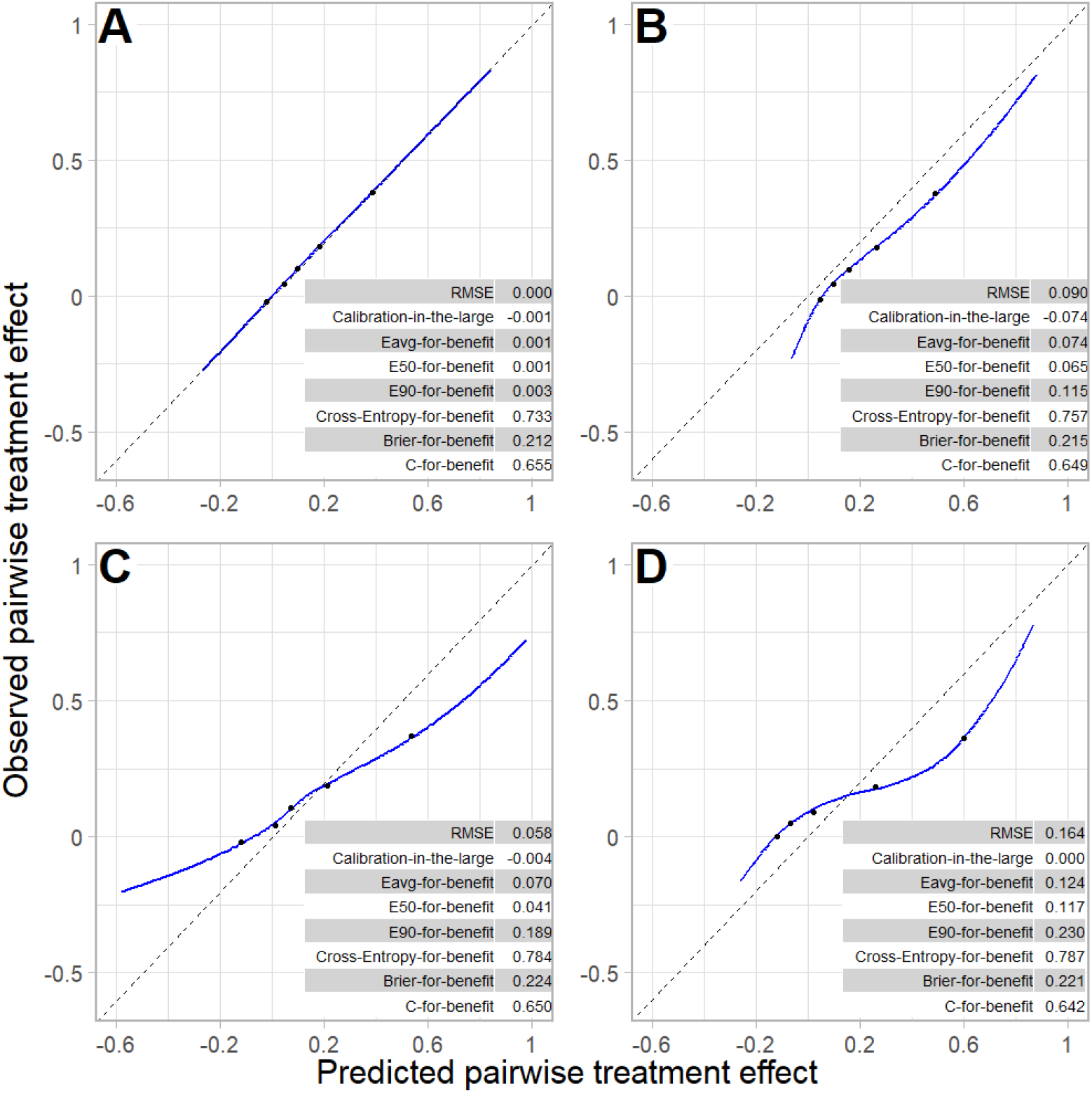
Calibration plots of pairwise treatment effect of simulated data from patients receiving lifestyle intervention. This Figure depicts observed versus predicted pairwise treatment effect by smoothed calibration curves (blue line) and quantiles of predicted pairwise treatment effect (black dots) of simulated data from the lifestyle intervention versus placebo treatment. Observed pairwise treatment effect was obtained by matching patients based on patient characteristics. Smoothed calibration curves were obtained by local regression of the observed pairwise treatment effect of matched patient pairs on predicted pairwise treatment effect of matched patient pairs. For prediction of individualized treatment effect, we used a risk-based “optimal model” (panel **A**) and three “perturbed models” that overestimate average treatment effect (panel **B**), risk heterogeneity (panel **C**), and treatment effect heterogeneity (panel **D**). The average treatment effect is 12.9, 20.4, 12.9 (after a correction of -0.14), and 12.9 (after a correction of 0.53), respectively.

The first “perturbed model” was designed to overestimate treatment effect of lifestyle intervention (RMSE=0.090), which was expressed graphically by the corresponding calibration curve lying below the 45-degree line, and numerically by suboptimal calibration metrics (calibration-in-the-large=-0.074, E_avg_-for-benefit=0.074, E_50_-for-benefit=0.065, E_90_-for-benefit=0.115, Figure 2B). The C-for-benefit expressed a slightly poorer ability to distinguish between patients with small and large treatment effects than the “optimal model” (C-for-benefit=0.649 versus 0.655). The cross-entropy-for-benefit and Brier-for-benefit also expressed poorer overall performance than the “optimal model” (cross-entropy-for-benefit=0.757 versus 0.733, Brier-for-benefit=0.215 versus 0.212, Figure 2A; 2B).

The second “perturbed model” was designed to overestimate risk heterogeneity of patients receiving lifestyle intervention (RMSE=0.058), which was expressed graphically by the corresponding calibration curve lying above the diagonal for low predicted pairwise treatment effect (underestimation of low treatment effect) and below the diagonal for high predicted pairwise treatment effect (overestimation of high treatment effect), and numerically by suboptimal calibration metrics (calibration-in-the-large=-0.004, E_avg_-for-benefit=0.070, E_50_-for-benefit=0.041, E_90_-for-benefit=0.189, Figure 2C). The C-for-benefit expressed a slightly poorer ability to distinguish between patients with small and large treatment effects than the “optimal model” (C-for-benefit=0.650 versus 0.655). The cross-entropy-for-benefit and Brier-for-benefit also expressed poorer overall performance than the “optimal model” (cross-entropy-for-benefit=0.784 versus 0.733, Brier-for-benefit=0.224 versus 0.212, Figure 2A; 2C).

The third “perturbed model” was designed to overestimate treatment effect heterogeneity of patients receiving lifestyle intervention (RMSE=0.164), which was expressed graphically by the corresponding calibration curve lying more extremely above the diagonal for low predicted pairwise treatment effect (underestimation of low treatment effect) and more extremely below the diagonal for high predicted pairwise treatment effect (overestimation of high treatment effect), and numerically by suboptimal calibration metrics (E_avg_-for-benefit=0.124, E_50_-for-benefit=0.117, E_90_-for-benefit=0.230, Figure 2D). The C-for-benefit expressed a slightly poorer ability to distinguish between patients with small and large treatment effects than the “optimal model” (C-for-benefit=0.642 versus 0.655, Figure 2D). The cross-entropy-for-benefit and Brier-for-benefit also expressed poorer overall performance than the “optimal model” (cross-entropy-for-benefit=0.787 versus 0.733, Brier-for-benefit=0.221 versus 0.212, Figure 2A; 2D).

The results from the simulations using the metformin treatment arm rather than the lifestyle intervention arm were similar (Figure 2; Additional file 6).

### 3.3 Case study

The differences in any of the performance measures between the risk model, the effect model, and the causal forest were not significantly different from zero in the 30 percent of patients who were in the test dataset (n=617; Additional file 3). Numerically, most calibration metrics of the effect model were better than that of the risk model (calibration-in-the-large=0.043 versus 0.052; E_avg_-for-benefit=0.050 versus 0.053; E_90_-for-benefit=0.123 versus 0.141, Figure 3A; 3B). Consequently, the overall performance of the effect model was numerically better than that of the risk model (cross-entropy-for-benefit=0.743 versus 0.747, Figure 3A; 3B), despite the numerically poorer discriminative ability of the effect model (C-for-benefit=0.663 versus 0.664, Figure 3A; 3B).

**Figure 3.**
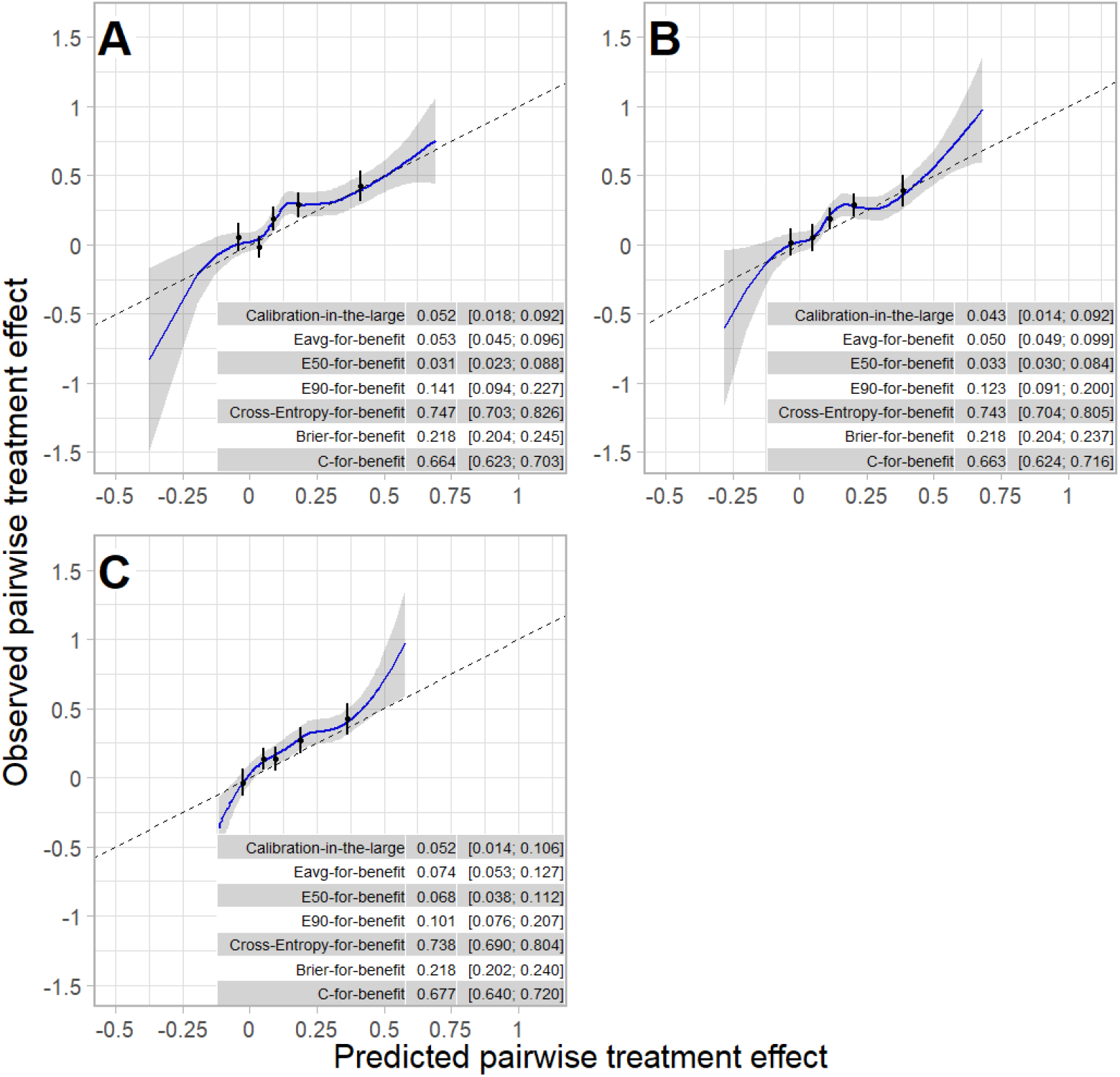
Calibration plot of pairwise treatment effect of simulated data from patients receiving lifestyle intervention. This Figure depicts observed versus predicted pairwise treatment effect by smoothed calibration curves (blue line with 95% confidence interval displayed by grey shaded area) and quantiles of predicted pairwise treatment effect (black dots) of lifestyle intervention versus placebo treatment. Observed pairwise treatment effect was obtained by matching patients based on patient characteristics. Smoothed calibration curves were obtained by local regression of the observed pairwise treatment effect of matched patient pairs on predicted pairwise treatment effect of matched patient pairs. For prediction of individualized treatment effect, we used: a risk modelling approach (panel **A**), a treatment effect modelling approach (panel **B**), and a causal forest (panel **C**). Confidence intervals around metric values were obtained using 100 bootstrap samples.

Central calibration metrics of the causal forest were numerically poorer than those of the risk model (E_avg_-for-benefit=0.074 versus 0.053; E_50_-for-benefit=0.068 versus 0.031, Figure 3A; 3C), but the causal forest resulted in less extreme miscalibration than the risk model (E_90_-for-benefit=0.101 versus 0.141, Figure 3A; 3C). Due to less extreme miscalibration and numerically better discriminative ability (C-for-benefit=0.677 versus 0.664, Figure 3A; 3C), the overall performance of the causal forest was numerically better than that of the risk model (cross-entropy-for-benefit=0.738 versus 0.747, Figure 3A; 3C).

## 4. Discussion

We extended the E-statistics, cross-entropy, and Brier score to quantify the quality of treatment effect predictions. The simulation study showed that the proposed metrics are useful for comparing models because the metrics of the data-generating model were consistently better than those of deliberately “perturbed models”. The case study illustrated the use of the proposed metrics in practice and showed that the calibration, discriminative ability, and overall performance of the three different models predicting treatment effect were not significantly different.

Similar to the previously proposed C-for-benefit, we defined observed pairwise treatment effect by the difference between outcomes in pairs of matched patients[6]. Matching patients based on predicted pairwise treatment effect would result in different patient pairs and consequently different observed pairwise treatment effect for each prediction model[6]. Therefore, we chose to match patients based on the Mahalanobis distance between patient characteristics resulting in the same observed pairwise treatment effect for each prediction model to allow for model comparison. The predicted pairwise treatment effect when matching patients by patient characteristics is more heterogeneous than the individual predicted treatment effect resulting from the model, which is more apparent in a smaller sample. We matched without replacement since the treatment arms were similar in size, but matching with replacement is more appropriate for samples with unbalanced treatment arms. Furthermore, we selected relevant patient characteristics based on clinical expertise and existing literature, but variable selection is more suitable in high-dimensionality data.

The case study is merely an illustration of the use of the performance metrics and not a framework for model selection or internal validation. The use of internal validation techniques other than split sampling is recommended for quantification of the performance of a model in similar settings, but that was outside the scope of this study[25]. The choice of the percentage of observations used for the training and test set was arbitrary. Furthermore, the proposed metrics in the training set will not be insightful when using models with penalization and honest tree building, because they will indicate by definition miscalibration in the training set (Additional file 7; 8).

The strength of our study is that we propose currently lacking performance metrics for models predicting treatment effect. Their actual values can be used to compare models predicting treatment effect. Furthermore, in future research updating strategies can be considered if our proposed calibration metrics indicate miscalibration of treatment effect predictions.

A limitation of this study is the limited sample size of the case study. In the simulation study, we showed that the performance metrics were able to distinguish between models for an artificially enlarged data set. However, in the case study, the confidence intervals of the performance metrics were overlapping. This phenomenon is inherent to treatment effect estimation. To obtain reasonable power, treatment effect analyses require a much larger sample size compared to when estimating an overall ATE[26]. The case study suggested that there is a trade-off between calibration and discrimination: better calibrated models were worse at discriminating between patients with small and large treatment effects, but due to the small sample size no strict conclusions can be drawn. Secondly, the performance metrics were developed for binary outcomes, which could be extended to continuous outcomes in future research. Notwithstanding these limitations, we conclude that the proposed metrics are useful to assess the calibration and overall performance of models predicting treatment effect.

## 5. Conclusions

We showed that the proposed metrics are useful to assess and compare the calibration and overall performance of models designed to predict treatment effect.

## Data Availability

Information on the process of obtaining the study dataset is available at the NIDDK Repository website (https://repository.niddk.nih.gov/studies/dpp/). The dataset can be obtained by submitting of a formal request to the NIDDK Repository.

https://github.com/CHMMaas/HTEPredictionMetrics

## 6. List of abbreviations

ATE: Average treatment effect
DPP: Diabetes Prevention Program
ICI: Integrated Calibration Index
IQR: Inter Quartile Range
RMSE: Root mean squared error

## DECLARATIONS

### Ethics approval and consent to participate

Not applicable

### Consent for publication

Not applicable

### Availability of data and materials

The dataset supporting the conclusions of this article is available in the NIDDK Repository website (https://repository.niddk.nih.gov/studies/dpp/). Project home page: https://github.com/CHMMaas/PaperPredictionMetrics.

### Authors’ contributions

**C.C.H.M. Maas**: Conceptualization, Methodology, Software, Formal analysis, Writing – Original Draft **D.M. Kent**: Conceptualization, Methodology, Writing – Review & Editing **M.C. Hughes**: Conceptualization, Writing – Review & Editing **R. Dekker**: Conceptualization, Writing – Review & Editing **H.F. Lingsma**: Conceptualization, Writing – Review & Editing **D. van Klaveren**: Conceptualization, Methodology, Writing – Review & Editing

## Acknowledgements

Not applicable

