## Supplementary Information for "Performance metrics for models designed to predict treatment effect"

**Additional file 1. Illustration of proposed metrics to assess models predicting treatment effect based on matching patients.**

|  | **Patient assigned to treatment** | | | | **Patient assigned to control treatment** | | | | **Matched pair** | | | | |
| --- | --- | --- | --- | --- | --- | --- | --- | --- | --- | --- | --- | --- | --- |
| **Matched patient pair (A)** | $\boldsymbol{p}_{\boldsymbol{0}}$ **(B)^*^** | $\boldsymbol{p}_{\boldsymbol{1}}$  **(C)^**^** | **Predicted treatment effect (D=B-C)** | **Observed outcome (E)** | $\boldsymbol{p}_{\boldsymbol{0}}$  **(F)^*^** | $\boldsymbol{p}_{\boldsymbol{1}}$  **(G)^**^** | **Predicted treatment effect (H=F-G)** | **Observed outcome (I)** | $\boldsymbol{p}_{\boldsymbol{0}}$ **(J=F)** | $\boldsymbol{p}_{\boldsymbol{1}}$ **(K=C)** | **Predicted pairwise treatment effect**  **(L=J-K)** | **Observed pairwise treatment effect (M=E-I)** | **LOESS curve (N)^***^** |
| 1 | 0.136 | 0.283 | -0.147 | 1 | 0.162 | 0.307 | -0.145 | 1 | 0.162 | 0.283 | -0.121 | 0 | 0 |
| 2 | 0.246 | 0.343 | -0.097 | 0 | 0.218 | 0.319 | -0.101 | 1 | 0.218 | 0.343 | -0.125 | -1 | -1 |
| 3 | 0.156 | 0.219 | -0.063 | 1 | 0.142 | 0.203 | -0.061 | 0 | 0.142 | 0.219 | -0.077 | 1 | 1 |
| 4 | 0.081 | 0.083 | 0.043 | 0 | 0.098 | 0.062 | 0.036 | 0 | 0.098 | 0.083 | 0.015 | 0 | 0 |
| 5 | 0.345 | 0.212 | 0.133 | 1 | 0.299 | 0.171 | 0.128 | 0 | 0.299 | 0.212 | 0.087 | 1 | 1 |
| 6 | 0.421 | 0.390 | 0.306 | 1 | 0.561 | 0.255 | 0.306 | 1 | 0.561 | 0.390 | 0.171 | 0 | 0 |

^*^$p_{0}=P\left( Y=1 | W=0 \right);$

^**^$p_{1}=P\left( Y=1 | W=1 \right);$

^***^$N=$ predict(loess($M \sim L$)), which results in the same values as the observed pairwise treatment effect (M), when rounded to three decimals, due to a small number of observations.

The calibration metrics:

Overall-calibration-for-benefit = abs(mean(M)-mean(N)) = 0

E­_avg_-for-benefit = mean(abs(L-N)) ≈ 0.529

E­_50_-for-benefit = median(abs(L-N)) ≈ 0.523

E­_90_-for-benefit = quantile(abs(L-N), 0.9) ≈ 0.995

The overall performance:

Cross-entropy-for-benefit $=-\frac{1}{n_{p}}\left[ I\left( M=1 \right)\cdot\log\left[ \left( 1-K \right)J \right]+I\left( M=0 \right)\log\left[ \left( 1-K \right)\left( 1-J \right)+K\cdot J \right]+ I\left( M=-1 \right)\log\left[ K\left( 1-J \right) \right] \right]\approx1.049$

Brier-for-benefit $=\frac{1}{2n_{p}}\left[ \left[ \left( 1-K \right)J-I\left( M=1 \right) \right]^{2}+\left[ \left( 1-K \right)\left( 1-J \right)+K\cdot J-I\left( M=0 \right) \right]^{2}+\left[ K\left( 1-J \right)-I\left( M=-1 \right) \right]^{2} \right] \approx0.321$

**Additional file 2. Derivation of the metrics measuring overall performance of models predicting treatment effect.**

*Derivation of the Brier-for-benefit*

The Brier-for-benefit is defined as

Brier-for-benefit$=\frac{1}{2n_{p}}\sum_{i=1}^{n_{p}} \sum_{c\in\left\{ -1,0,1 \right\}} \left( P\left( \tau_{i}=c \right)-I\left( \tau_{i}=c \right) \right)^{2},$

where $n_{p}$ indicates the number of pairs, $\tau_{i}$ indicates the observed pairwise treatment effect in a matched pair $i$, $I\left( \tau_{i}=c \right)$ is an indicator function returning one when the observed pairwise treatment effect of matched pair $i$ $\left( \tau_{i} \right)$ is equal to class $c$, and $P(\tau_{i}=c)$ indicates the probability that the observed pairwise treatment effect of matched pair $i$ is equal to class $c$. The Brier score is divided by two to ensure that it lies between zero and one because in the worst-case scenario you give the highest prediction (one) for the wrong class, which would give a Brier score of two. Equivalently,

Brier-for-benefit$=\frac{1}{2n_{p}}\sum_{i=1}^{n_{p}} \left[ \left( P\left( \tau_{i}=1 \right)-I\left( \tau_{i}=1 \right) \right)^{2}+\left( P\left( \tau_{i}=0 \right)-I\left( \tau_{i}=0 \right) \right)^{2}+\left( P\left( \tau_{i}=-1 \right)-I\left( \tau_{i}=-1 \right) \right)^{2} \right]$

Since matched patient pairs are independent, it holds that

$$P\left( \tau_{i}=1 \right)=P\left( Y_{i}\left( 1 \right)=0, Y_{i}\left( 0 \right)=1 \right)$$

$$=P\left( Y_{i}\left( 1 \right)=0 \right)P\left( Y_{i}\left( 0 \right)=1 \right)$$

$$=\left( 1-p_{i,1} \right)p_{i,0}$$

$$P\left( \tau_{i}=0 \right)=P\left( \left( Y_{i}\left( 1 \right)=0, Y_{i}\left( 0 \right)=0 \right)\cap\left( Y_{i}\left( 1 \right)=1, Y_{i}\left( 0 \right)=1 \right) \right)$$

$$=P\left( Y_{i}\left( 1 \right)=0 \right)P\left( Y_{i}\left( 0 \right)=0 \right)+P\left( Y_{i}\left( 1 \right)=1 \right)P\left( Y_{i}\left( 0 \right)=1 \right)$$

$$=\left( 1-p_{i,1} \right)\left( 1-p_{i,0} \right)+p_{i,1}p_{i, 0}$$

$$P\left( \tau_{i}=-1 \right)=P\left( Y_{i}\left( 1 \right)=1, Y_{i}\left( 0 \right)=0 \right)$$

$$=P\left( Y_{i}\left( 1 \right)=1 \right)P\left( Y_{i}\left( 0 \right)=0 \right)$$

$$=p_{i,1}\left( 1-p_{i,0} \right),$$

where $Y_{i}\left( W_{i} \right)=\left\{ \begin{aligned} Y_{i}\left( 0 \right) \mathrm{if} W_{i}=0 \\ Y_{i}\left( 1 \right) \mathrm{if} W_{i}=1 \end{aligned} \right.$ with $Y_{i}$ indicates the potential outcome for patient $i$ and $W_{i}$ indicates the binary indicator for treatment, and the outcome probabilities conditional on treatment

$$p_{i,1}=P\left( Y_{i}\left( 1 \right)=1 \right)=P\left( Y_{i}=1 | W_{i}=1 \right)$$

$$p_{i,0}=P\left( Y_{i}\left( 0 \right)=1 \right)=P\left( Y_{i}=1 | W_{i}=0 \right).$$

As a result, the Brier-for-benefit can be expressed as

Brier-for-benefit$=\frac{1}{2n_{p}}\sum_{i=1}^{n_{p}} \left( \left( 1-p_{i,1} \right)p_{i,0}-I\left( \tau_{i}=1 \right) \right)^{2}$

$$=+\frac{1}{2n_{p}}\sum_{i=1}^{n_{p}} \left( \left( 1-p_{i,1} \right)\left( 1-p_{i,0} \right)+p_{i,1}p_{i,0}-I\left( \tau_{i}=0 \right) \right)^{2}$$

$$=+\frac{1}{2n_{p}}\sum_{i=1}^{n_{p}} \left( p_{i,1}\left( 1-p_{i,0} \right)-I\left( \tau_{i}=-1 \right) \right)^{2}.$$

*Derivation of the cross-entropy-for-benefit*

Similarly, the cross-entropy-for-benefit is defined as

Cross-entropy-for-benefit $=-\frac{1}{n_{p}}\cdot\sum_{i=1}^{n_{p}} \sum_{c\in\left\{ -1,0,1 \right\}} I\left( \tau_{i}=c \right)\log\left[ P\left( \tau_{i}=c \right) \right]$

$$=-\frac{1}{n_{p}}\cdot\sum_{i=1}^{n_{p}} I\left( \tau_{i}=1 \right)\log\left[ \left( 1-p_{i,1} \right)p_{i,0} \right]$$

$$-\frac{1}{n_{p}}\cdot\sum_{i=1}^{n_{p}} I\left( \tau_{i}=0 \right)\log\left[ \left( 1-p_{i,1} \right)\left( 1-p_{i,0} \right)+p_{i,1}p_{i,0} \right]$$

$$-\frac{1}{n_{p}}\cdot\sum_{i=1}^{n_{p}} I\left( \tau_{i}=-1 \right)\log\left[ p_{i,1}\left( 1-p_{i,0} \right) \right].$$

*Outcome probabilities of the causal forest*

Of note, the outcome probabilities conditional on treatment $p_{i, 0}$ and $p_{i,1}$ probabilities of the causal forest are obtained by

$$p_{i,0}=E\left[ Y | X,W=0 \right]=\hat{m}\left( X \right)-\hat{e}\left( X \right)\hat{\tau}(X)$$

$$p_{i,1}=E\left[ Y | X, W=1 \right]=\hat{m}\left( X \right)+\left( 1-\hat{e}\left( X \right) \right)\hat{\tau}\left( X \right),$$

with $\hat{m}\left( X \right)=E[Y|X=x]$ and $\hat{e}\left( X \right)=E\left[ W | X=x \right]$ indicate the outcomes of two random forests, and $\hat{\tau}\left( X \right)=E\left[ Y_{i}\left( 0 \right)-Y_{i}\left( 1 \right) | X=x \right]$ indicates the treatment effect outcomes.

### **Additional file 3. Characteristics of patients in the Diabetes Prevention Program receiving lifestyle intervention, metformin, or placebo treatment.**

|  | Total | | |  | Lifestyle | |  | Metformin | |  | Placebo | |
| --- | --- | --- | --- | --- | --- | --- | --- | --- | --- | --- | --- | --- |
|  | N | % | Missing |  | N | % |  | N | % |  | N | % |
| Sample size | 3081 |  |  |  | 1024 |  |  | 1027 |  |  | 1030 |  |
| Diabetes | 655 | 21.3 |  |  | 148 | 4.8 |  | 215 | 7.0 |  | 292 | 9.5 |
| Female | 2053 | 66.6 |  |  | 685 | 22.2 |  | 669 | 21.7 |  | 699 | 22.7 |
| Ethnicity |  |  |  |  |  |  |  |  |  |  |  |  |
| Black | 644 | 20.9 |  |  | 204 | 6.6 |  | 221 | 7.2 |  | 219 | 7.1 |
| Hispanic | 508 | 16.5 |  |  | 178 | 5.8 |  | 162 | 5.3 |  | 168 | 5.5 |
| History of high blood glucose | 614 | 19.9 |  |  | 206 | 6.7 |  | 192 | 6.2 |  | 216 | 7.0 |
| Family history of diabetes | 2127 | 69.0 | 2 |  | 713 | 23.1 |  | 699 | 22.7 |  | 715 | 23.2 |
| Smoking | 216 | 7.0 |  |  | 67 | 2.2 |  | 69 | 2.2 |  | 80 | 2.6 |
| Hypertension | 835 | 27.1 |  |  | 286 | 9.3 |  | 267 | 8.7 |  | 282 | 9.2 |
| Gestational diabetes mellitus | 321 | 10.4 | 1 |  | 108 | 3.5 |  | 106 | 3.4 |  | 107 | 3.5 |
| Age | 52 | [42; 57] |  |  | 47 | [42; 57] |  | 52 | [42; 57] |  | 47 | [42; 57] |
| BMI | 33 | [29; 37] |  |  | 33 | [29; 37] |  | 33 | [29; 37] |  | 33 | [29; 37] |
| Triglycerides | 141 | [99; 201] | 5 |  | 138 | [97; 200] |  | 137 | [98; 195] |  | 147 | [104; 207.5] |
| Haemoglobin $\boldsymbol{A}_{\boldsymbol{1}\boldsymbol{c}}$ | 5.9 | [5.6; 6.2] | 8 |  | 5.9 | [5.6; 6.2] |  | 5.9 | [5.6; 6.2] |  | 5.9 | [5.6; 6.2] |
| Fasting blood sugar | 105 | [101; 112] |  |  | 105 | [101; 112] |  | 105 | [100; 112] |  | 106 | [101; 112] |

**Additional file 4. The probability of diabetes for each level of the prognostic index on the log odds scale when not treating (blue) and treating patients (red) with lifestyle intervention.** This Figure displays the “optimal model” in panel **A**, and three “perturbed models” that overestimate average treatment effect (panel **B**), risk heterogeneity (panel **C**), and treatment effect heterogeneity (panel **D**).

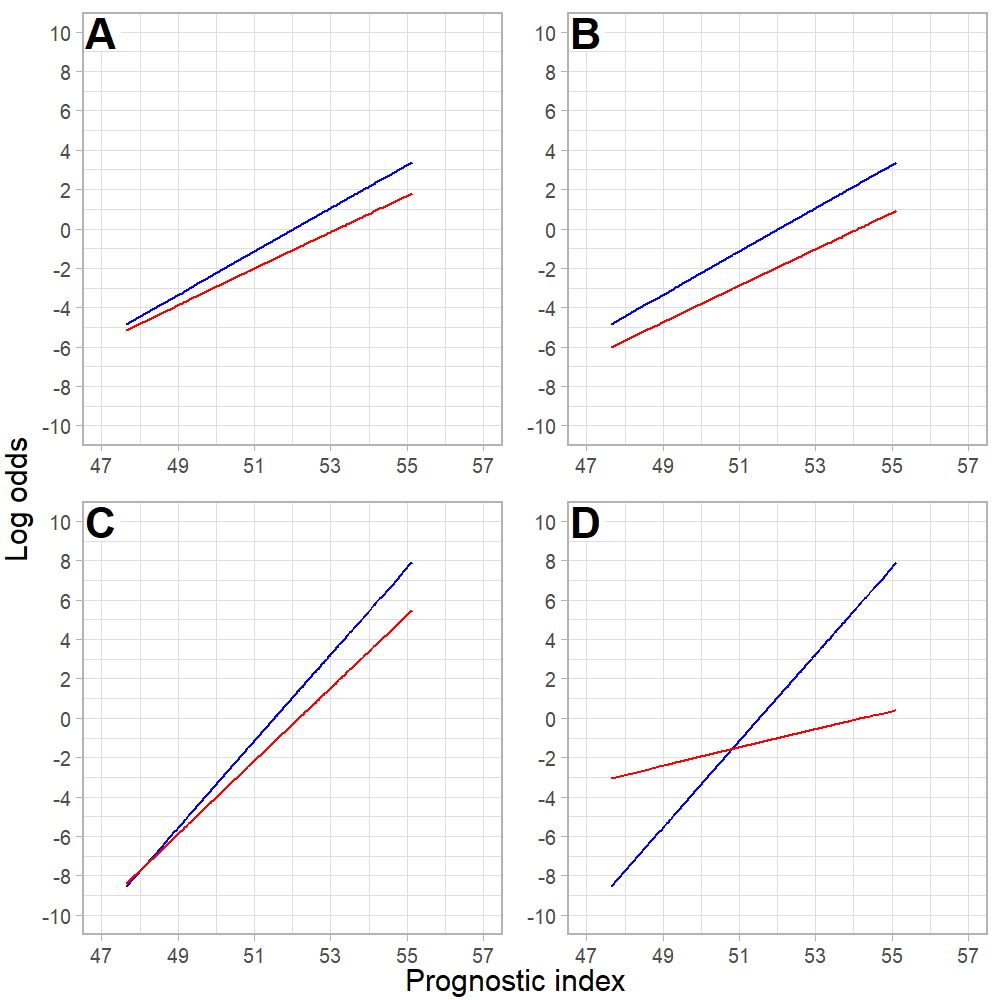

**Additional file 5. The probability of diabetes for each level of the prognostic index on the log odds scale when not treating (blue) and treating patients (red) with metformin.** This Figure displays the “optimal model” in panel **A**, and three “perturbed models” that overestimate average treatment effect (panel **B**), risk heterogeneity (panel **C**), and treatment effect heterogeneity (panel **D**).

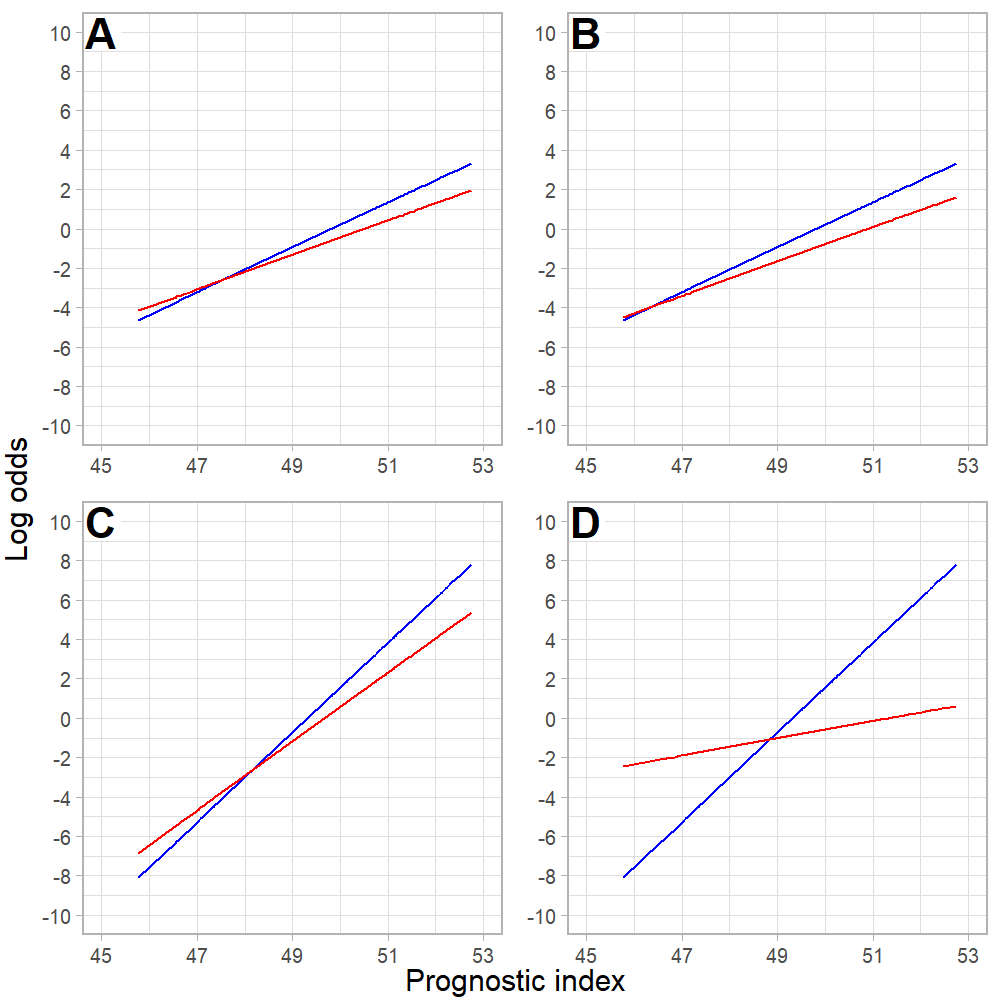

**Additional file 6. Calibration plots of pairwise treatment effect of simulated data from patients receiving metformin intervention.** This Figure depicts observed versus predicted pairwise treatment effect by smoothed calibration curves (blue line) and quantiles of predicted pairwise treatment effect (black dots) of simulated data from the metformin versus placebo treatment. Observed pairwise treatment effect was obtained by matching patients based on patient characteristics. Smoothed calibration curves were obtained by local regression of the observed pairwise treatment effect of matched patient pairs on predicted pairwise treatment effect of matched patient pairs. For prediction of individualized treatment effect, we used a risk-based “optimal model” (panel **A**) and three “perturbed models” that overestimate average treatment effect (panel **B**), risk heterogeneity (panel **C**), and treatment effect heterogeneity (panel **D**). The average treatment effect is 6.5, 11.1, 6.5 (after a correction of -0.02), and 6.5 (after a correction of 0.375), respectively.
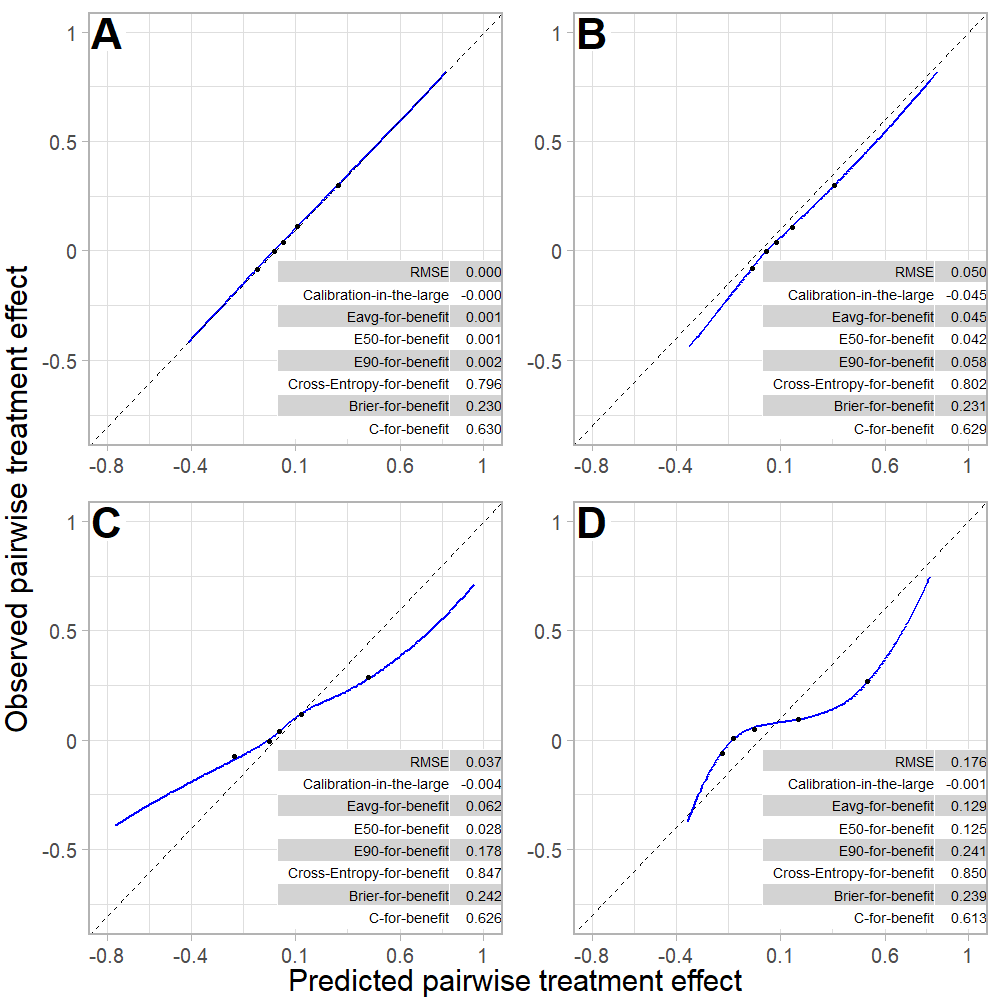

**Additional file 7. Calibration plot of pairwise treatment effect of training and test data of lifestyle intervention.** This Figure depicts observed versus predicted pairwise treatment effect by smoothed calibration curves (blue line with 95% confidence interval displayed by grey shaded area) and quarters of predicted pairwise treatment effect (black dots) of lifestyle intervention versus placebo treatment. Observed pairwise treatment effect was obtained by matching patients based on patient characteristics. Smoothed calibration curves were obtained by local regression of the observed pairwise treatment effect of matched patient pairs on predicted pairwise treatment effect of matched patient pairs. For prediction of treatment effect, we used: a risk modelling approach (panel **A**; **B**), a treatment effect modelling approach (panel **C**; **D**), and a causal forest (panel **E**; **F**). The models are trained on 70 percent of the data (panel **A**; **C**; **E**) and evaluated on the other 30 percent of the data (**B**; **D**; **F**). Confidence intervals around the metric values were obtained using 100 bootstrap samples.

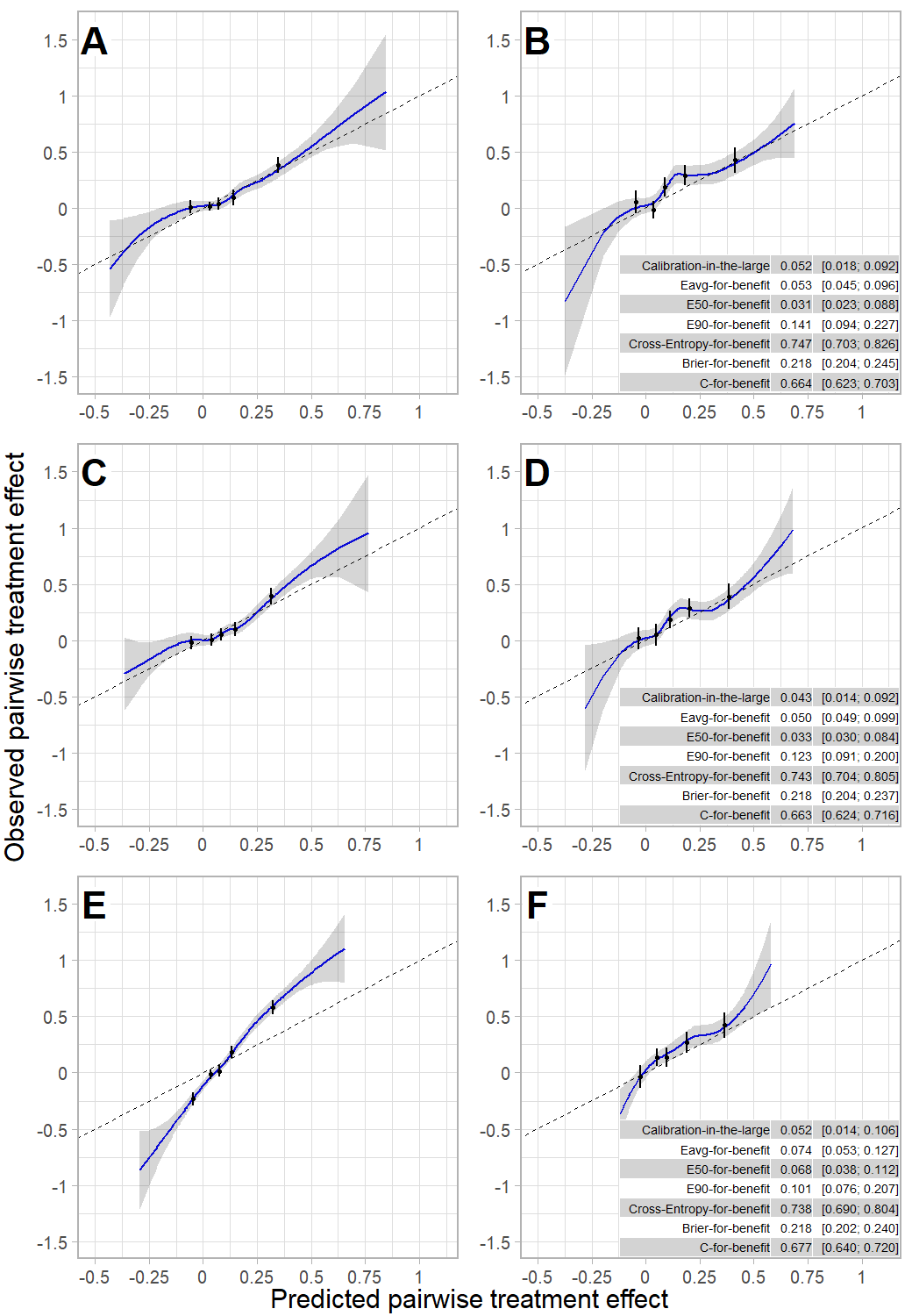

**Additional file 8. Calibration plot of pairwise treatment effect of training and test data of metformin intervention.** This Figure depicts observed versus predicted pairwise treatment effect by smoothed calibration curves (blue line with 95% confidence interval displayed by grey shaded area) and quarters of predicted pairwise treatment effect (black dots) of metformin versus placebo treatment. Observed pairwise treatment effect was obtained by matching patients based on patient characteristics. Smoothed calibration curves were obtained by local regression of the observed pairwise treatment effect of matched patient pairs on predicted pairwise treatment effect of matched patient pairs. For prediction of treatment effect, we used: a risk modelling approach (panel **A**; **B**), a treatment effect modelling approach (panel **C**; **D**), and a causal forest (panel **E**; **F**). The models are trained on 70 percent of the data (panel **A**; **C**; **E**) and evaluated on the other 30 percent of the data (**B**; **D**; **F**). Confidence intervals around the metric values were obtained using 100 bootstrap samples.

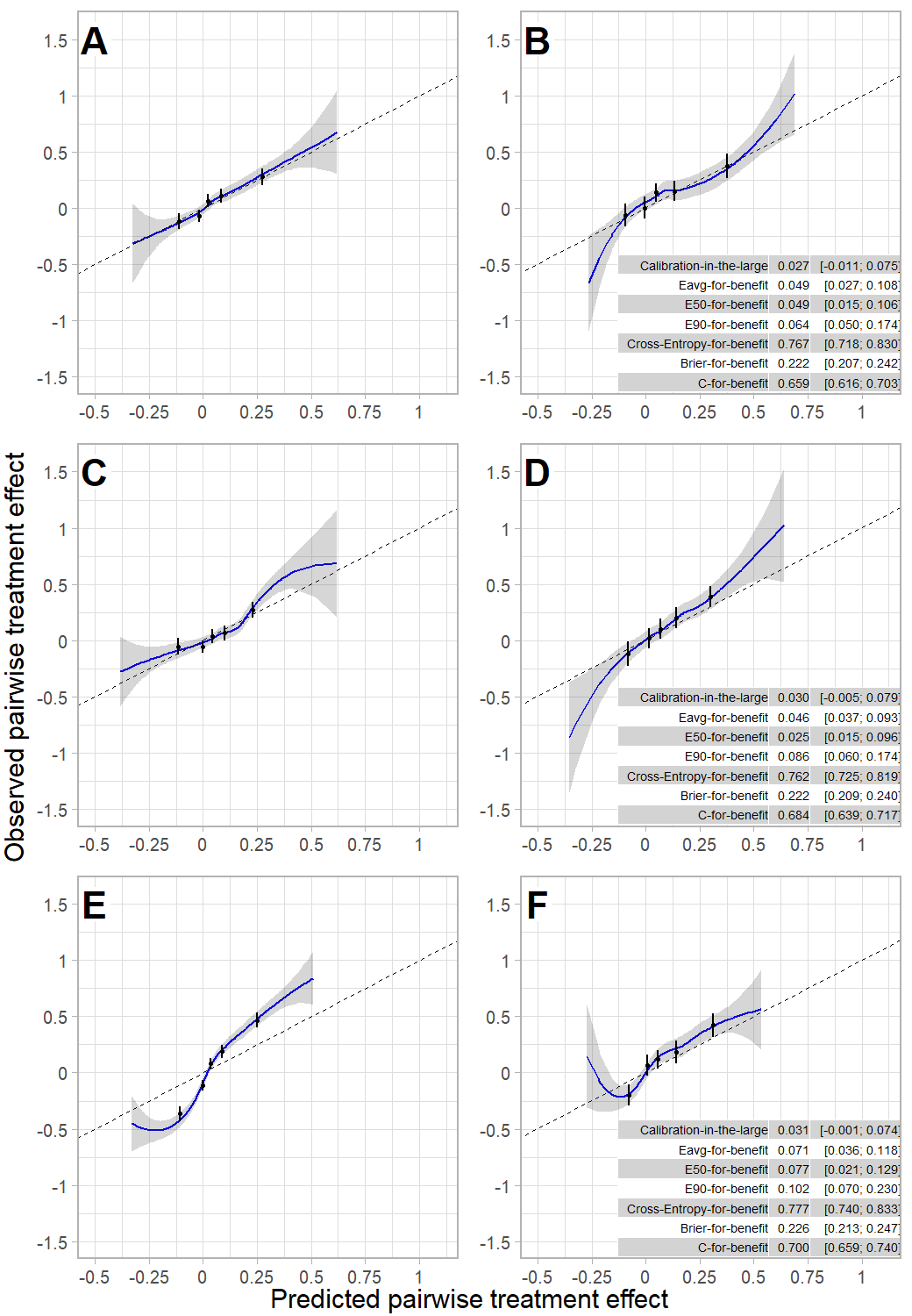
